# Law, Criminalisation and HIV in the World: Have countries that criminalise achieved more or less successful AIDS pandemic response?

**DOI:** 10.1101/2021.06.04.21258360

**Authors:** Matthew M. Kavanagh, Schadrac C. Agbla, Mara Pillinger, Marissa Joy, Alaina Case, Ngozi Erondu, Kashish Aneja, Taavi Erkkola, Ellie Graeden

## Abstract

How does the use of criminal law affect disease-fighting efforts, particularly in a pandemic? This longstanding question for governments around the world is felt acutely in the context of the COVID-19 and HIV pandemics. Many countries have laws and policies that criminalise behaviours, making same-sex relationships, illicit drug use, and sex work illegal. Meanwhile, some countries have enshrined gender- and rights-protective institutions in law. Under the global AIDS strategy of the last five years, national AIDS response efforts in countries have focused on reaching people living with HIV with testing and antiretroviral treatment to suppress the HIV virus, preventing mortality and HIV transmission. At the end of this 5-year push, this article provides an ecological analysis of whether those countries with criminalising legal environments achieved more or less success. In countries where same-sex relationships were fully criminalised, the portion of people living with HIV who knew their status was 11% lower and viral suppression rates were 8% lower. Under sex-work criminalization, the rate of people living with HIV who knew their status was 10% lower and viral suppression 6% lower. Drug use criminalisation was associated with 14% lower knowledge of status and viral. On the other hand in countries with laws advancing non-discrimination, human rights institutions, and gender-based violence response, HIV services indicators were significantly better. This ecological evidence on the relationships between the legal environment and successful HIV response provides support for a strategy that includes a focus on law reform to achieve goals missed in 2020.

**Summary Box:**

- Countries around the world, across economic and geographic boundaries, have taken different approaches to the application of criminal law to same-sex sex, sex work, and drug use—with most taking a partially or fully criminalising legal approach in one or more of these areas.
- In tackling the AIDS pandemic, globally agreed goals for 2020 focused on ensuring most people living with HIV were aware of their HIV status and had suppressed the HIV virus through effective antiretroviral treatment.
- The AIDS response in countries that criminalised was less successful than those that did not—achieving significantly lower levels of HIV status knowledge and HIV viral suppression.
- Countries with clear laws advancing non-discrimination, human rights institutions, and gender-based violence response had better knowledge of HIV status and viral suppression rates.
- This analysis suggests a new global AIDS strategy that includes a focus on law reform may hold promise in achieving goals that were missed in 2020.

## Introduction

Pandemic diseases like HIV present a significant challenge for public health. With neither a cure nor a vaccine available, halting the pandemic requires a mix of biomedical and social interventions aimed at averting death and reducing new infections. The legal and policy environment in a given country context shapes vulnerability to disease as well as roll out of, and participation in, interventions, and has been identified as a determinant of health outcomes.^1–3^

During the last decade biomedical science has progressed rapidly, with breakthroughs in prevention and treatment.^4,5^ Yet the global HIV response has seen highly differential success between countries. While AIDS deaths and new infections fell, according to UN estimates only 26 countries are on track to achieve the global goal of a 90% reduction in AIDS-related mortality by 2030 and only 23 countries are on track to reduce new HIV infections by 90%.^6^

Globally, at the end of 2019, an estimated 81% of people living with HIV (PLHIV) knew their HIV status.^7^ By June 2020, 26 million PLHIV were accessing treatment—a number that has more than tripled since 2010. In total, 59% of all PLHIV knew their status, were on antiretroviral treatment, and had suppressed virus, which improves health and prevents both mortality and HIV transmission.

Countries across different regions, income levels, and epidemic profiles that have seen success. In Thailand, for example, new infections have fallen by two thirds since 2010 as over 90% of all PLHIV came to know their status and 78% achieved viral suppression. Germany had similar levels of viral suppression to Thailand while South Africa, with the largest HIV epidemic in the world, had an even higher 92% knowledge of HIV status but lower viral suppression as it struggled to keep people on treatment. The United States, the world’s wealthiest nation, had lower rates than all three. And at the other end of the spectrum, in more than a quarter of countries less than 50% of PLHIV had suppressed virus in 2019.^6^ These include wealthier countries like Russia and lower middle income countries like Pakistan, where only an estimated 21% of PLHIV know their status and new infections are rising. In Jamaica and Angola, AIDS deaths are rising amidst low rates of viral suppression; just 35% of HIV-positive Jamaicans have suppressed virus.

Law is one of several epidemiologic, social, economic, and political factors driving differential success. As the Global Commission on HIV and the Law found, “the legal environment can play a powerful role in the well-being of people living with HIV and those vulnerable to HIV.”^8^ Law is particularly powerful when it comes to key populations experiencing higher HIV rates and lower access to services, including gay men and other men who have sex with men, sex workers, transgender people, people who inject drugs.^9^ Laws that subject key populations to arrest, prosecution, and imprisonment for engaging in behaviours governments deem undesirable are widespread. Meanwhile, many countries have what we call rights- and gender-protective laws which establish independent rights institutions, combat gender-based violence, and outlaw discrimination.

The UN-adopted Global AIDS Strategy ending in 2021 centered 90-90-90 targets^1^ on HIV testing and treatment, aiming for 90% of PLHIV to know their status a total of 73% of all PLHIV to have suppressed virus by 2020.^10^ A new strategy for the 2022-2027 period specifically focuses on removal of criminalising laws alongside expansion of efforts to combat stigma, gender-based violence and human rights violations.^11^

This paper explores whether countries that pursued a criminalisation approach achieved better or worse outcomes under the 90-90-90 targets than those with a less criminalised environment. We further ask whether countries that adopted rights- and gender-protective laws and policies ended this period with better HIV outcomes or not. We do so using a new dataset that has coded HIV-related laws and policies for all 194 UN member countries to enable cross-national comparative analysis.^12^

### Criminal Laws, Rights & Gender Institutions, and HIV

Since long before HIV, through to the present, governments have argued that criminalising disfavored behaviour like same-sex sex, sex work, and drug use helps prevent disease and change behaviour.^13–15^ Though scholars seldom go that far, some have argued against decriminalisation on the grounds it could have negative health implications.^16^ A wider normative, theoretical, and empirical literature finds criminalisation has a negative effect on HIV services and on health outcomes for key populations.^17–22,9^ Empirical work in both high-income and lower-income settings has included population-specific qualitative studies in specific countries or regions,^23–26^ quantitative modeling,^27^ ecological studies in particular regions,^28,29^ and increasingly sophisticated quantitative efforts to address causality in multi-country samples.^30^

Here we explore the question of whether criminalising countries saw better or worse HIV outcomes at the end of the last global AIDS strategy using a new dataset of laws and policies for all 194 UN member countries. The *HIV Policy Lab* dataset provides cross-nationally comparative coded data.^12^ This enables us to look three aspects in global perspective: same-sex, sex work, and drug use/possession criminalisation. We explore all three simultaneously, and in combination, as well as rights- and gender-protective laws. Data is gathered from legal texts, analysis of official reporting to the UN, and a meta-analysis of additional public sources, as described in Kavanagh et al.^12^

Around the world same-sex, sex work, and drug use criminalisation is widespread but varied, cutting across income and geography. As of 2020 only 20% of countries fully criminalise all three aspects explored here but every country at least partially criminalises one; 21% and 54% fully criminalise one or two respectively. Meanwhile, 23% of countries have nondiscrimination protections that cover sexual orientation, gender identity, and HIV status while 39% have independent human rights institutions and 75% have enforceable gender-based violence laws.

### Assessing Laws and Pandemic Response Progress

The 90-90-90 targets, set in 2014 to be achieved by 2020, give us one clear, globally-agreed metric with which to compare the success of national AIDS responses. Knowledge of status is a critical entry-point for differential biomedical and behavioural options for prevention as well as treatment. Viral suppression is a key indicator of the accessibility, acceptability, and quality of HIV services. Criminalisation has been linked to a variety of factors, from mistrust of authorities to stigma from providers, that leads to avoidance of medical services, avoidance of diagnosis, and challenges starting and staying on HIV treatment.^19,24,30^ Whether those resulted in measurable global differences in the overall success of countries’ AIDS response—particularly during this period of global focus on 90-90-90 targets—is not clear.

We use a cross-sectional dataset to compare the relative success of criminalising countries to those with a less punitive legal approach. Our outcome variables for each country are the percent of all PLHIV who know their status and the percent of all PLHIV who are virally suppressed, both from UNAIDS data released July 2020.^7^

We test four variables of interest related to criminalisation from the HIV Policy Lab against these HIV service outcomes. Same-sex criminalisation codes whether the laws of a given country criminalise consensual same-sex sex acts and whether the country actively prosecutes people under these laws. The variable on sex work codes whether national law criminalises the buying, selling, and organizing of sex work. The drug use variable codes whether national laws criminalises the use or possession of small amounts of drugs, including opioids, for personal consumption. Each of these variables is coded on a three part scale of criminalised, partially criminalised, or not criminalised. This range is important since, while seeking the benefits of quantitative coding, we are conscious of work on the complex relationship between criminalisation and health. We drop the few observations where countries laws meaningfully changed in this period. We also test a fourth variable that combines all three of these into overall criminalisation of key population-linked behaviour. Full descriptive statistics and exploration of the data is available at HIVPolicyLab.org.

We use fractional logistic regression models with robust standard errors to investigate the association between HIV services outcomes and explanatory variables of interest. Fractional logistic regressions with robust standard error are suitable for variables whose values vary from 0 to 1 or as percentages like in our setting and provide consistent estimates.^31^ We subsequently estimate the marginal average effects interpreted as the amount of change in the outcome induced by changing the law status from “not adopted” to “fully adopted”. When explanatory variables are log-transformed, the marginal average effects are interpreted as the amount of change in the outcome induced by one percent increase in the log-transformed explanatory variable.

We also explore whether countries with rights- and gender-protective laws like those described in the new Global AIDS Strategy have achieved better HIV-outcomes during the previous strategy period. Here we test laws on: protections from discrimination based on sexual orientation, gender identity, and HIV status; an independent national human rights institution to which PLHIV and key populations to report abuse and discrimination that meets the conditions set out in the UN Paris Principles;^32^ and explicit gender-based violence laws with enforceable penalties.

In the multivariate analysis for each variable we control for basic confounders. These include relative wealth and spending on health in the country—log health expenditure per capita, 2019^33^— which has a clear theoretical connection to success in HIV service provision, and a significant association in our data. We also control for the relative size of the HIV epidemic in a country to account for the distinct but significant challenges reaching high coverage in both high-prevalence and low-prevalence settings, using a baseline 2014 prevalence,^34^ the year the 90-90-90 goals were set.

While we cannot make causal inferences from this data, we can understand the basic question: did countries with criminalising or rights/gender protecting laws achieve better, worse, or roughly the same outcomes over this period compared to those who did not?

### Evidence on the AIDS Response Under Criminalisation and Under Rights-Protection

In countries with criminalised legal environments, a smaller portion of people living with HIV knew their HIV status and had suppressed virus compared to countries with less criminalising laws. These relationships are shown in Figure 1, with coefficients and details in Table 1. Countries that criminalise same-sex relationships had an unadjusted average of 11.3% (p=0.002) lower knowledge of HIV status among people living with HIV. In countries that criminalise sex work 10.2% (p=0.06) less of those living with HIV know their status and where drug use is criminalised it is 14.2% (p=0.008) lower. Taken together in our composite measure, the model shows that if trends held across countries, a theoretical country with no criminalisation across these all three areas would have as much as 24.0% higher knowledge of status than one with a fully criminalised environment on all three (p=0.001), noting a coefficient of 8.0 representing a one-unit increase). After controlling for HIV prevalence and health expenditure per capita, same sex criminalisation retained significance at the 10% level, while the other findings remained substantial but lost statistical significance—which was not surprising for cross-sectional data with a relatively small N compared with factors like prevalence highly correlated to our outcome variables.

**Figure 1:**
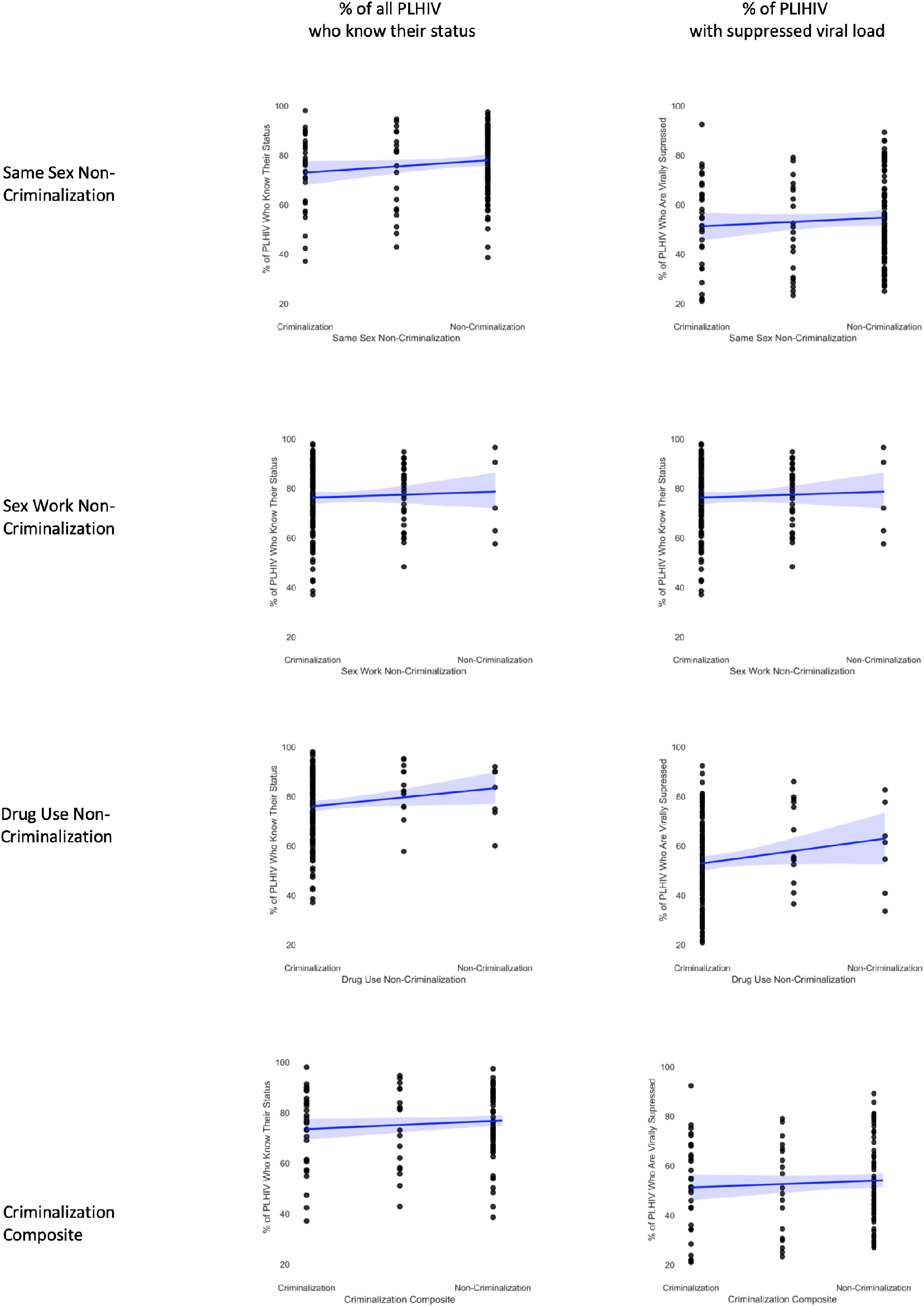
HIV Services Outcomes Under Punitive Laws.

Countries with criminalised legal environments had lower rates of viral suppression compared to those with non-criminalised environments. Rates of viral suppression were, in an unadjusted average, 8.1% (p=0.04) lower in countries that criminalise same-sex relationships, 5.9% (p=0.27) lower under sex-work criminalisation, and 14.5% (p=0.02) lower when drug use was criminalised. Taken together the difference between a theoretically fully criminalised and fully non-criminalised country would be as much as an 18.3% (p=0.01) difference in viral suppression.

Further analysis is worthwhile in the future when a longitudinal dataset with a larger sample size can be created and where broader epidemiologic measures like incidence changes can be explored.

A higher portion of people living with HIV knew their HIV status in countries with non-discrimination protections (9.7 %, p=<0.001), independent human rights institutions (3.0%, p=0.08), or explicit gender-based violence laws (15.9%, p=0.001). Figure 2 shows these relationships. Higher viral suppression rates were also significantly associated with non-discrimination protections (10.7%, p=<0.001), independent human rights institutions (3.3%, p=0.06), and gender-based violence laws (16.2%, p=<0.001). Non-discrimination and gender-based violence laws were significant even after controlling for prevalence and spending.

**Figure 2:**
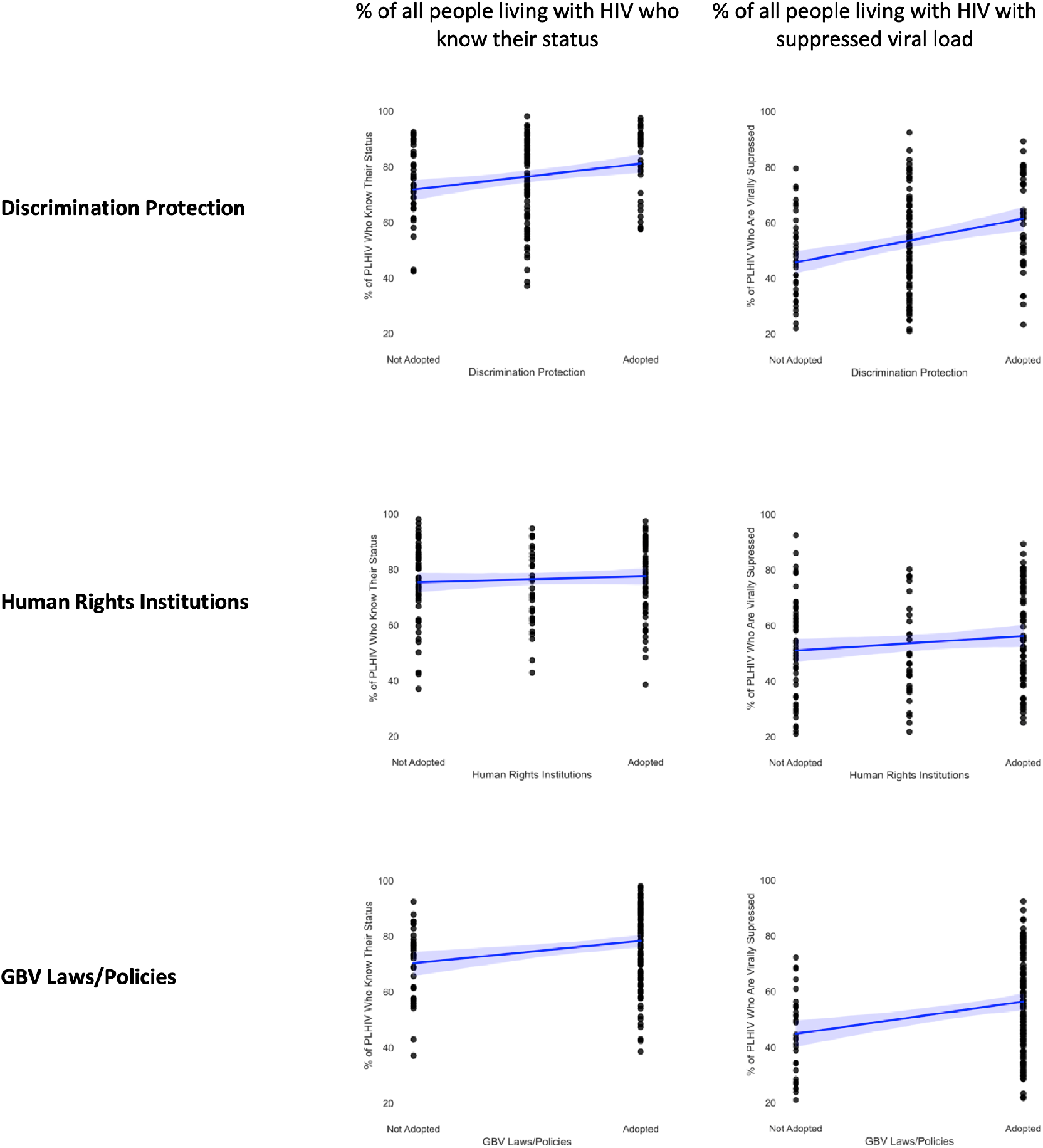
HIV Services Outcomes Under Rights- and Gender-Protective Laws.

There are a number of important limitations to this analysis. First, this is an ecological study and is not sufficient to identify causal mechanisms. Ecological fallacies and confounding factors cannot be discounted. One strength of this analysis is a unique global dataset addressing countries in all regions and income levels—allowing us to avoid common problems of systematically missing but theoretically important countries that challenges cross-national social science.^35^ Yet while multi-level data would be desirable to study HIV outcomes, no such global dataset exists.^36^ In addition, we face all of the common problems of measuring law, including that our tripartite coding rules cannot capture enforcement or subtle differences between countries that may be meaningful. As such the causal effect of criminalisation and protective laws remains an important topic of future research.

## Conclusion

At the end of a 5-year strategy in which countries around the world focused their AIDS response on reaching people living with HIV with testing and treatment services, countries that have adopted a criminalising approach to key populations saw less success than those that chose not to criminalise. Where same-sex relationships, sex work, and drug use were criminalised, a smaller portion of PLHIV knew their status and fewer had suppressed virus. Meanwhile, there is a positive association between these HIV outcomes and the adoption of laws that advance non-discrimination, human rights institutions, and responses to gender-based violence. These relationships were observable by using a global database of comparable cross-national data about laws and policies. As this resource grows and longitudinal data become available, studies with greater causal identification will become increasingly possible.

In the last year, laws went into effect in Angola removing criminalisation of same-sex sex while enacting non-discrimination protections on the basis of sexual orientation; in the U.S. state of Oregon decriminalising personal drug possession; and in Northern Territory Australia decriminalising sex work. This study suggests these may well be positive moves for public health, ripe for further study.

This study finds little support for the argument that criminalising behaviour among marginalized people in a pandemic results in positive outcomes. This is an important point of departure for future analyses in the era not just of HIV but the COVID-19 pandemic, tuberculosis, and many other contexts.

## Data Availability

All data are available online at www.HIVPolicyLab.org and https://aidsinfo.unaids.org/

https://aidsinfo.unaids.org/

## Footnotes

1 90-90-90 targets are 90% of all people living with HIV know their HIV status; 90% of those people are on sustained antiretroviral treatment; and 90% of those on treatment have viral suppression.

## References

1. Grover A. Report of the Special Rapporteur on the Right of Everyone to the Enjoyment of the Highest Attainable Standard of Physical and Mental Health. Geneva: United Nations Office of the High Commissioner; 2011.

2. Avafia T, Konstantinov B, Esom K, Sanjuan JR, Schleifer R. A rights-based response to COVID-19: Lessons learned from HIV and TB epidemics. Health and Human Rights Journal. 2020;

3. Gostin LO, Monahan JT, Kaldor J, DeBartolo M, Friedman EA, Gottschalk K, et al. The legal determinants of health: harnessing the power of law for global health and sustainable development. The Lancet. 2019;393(10183):1857–910.

4. Abdool Karim SS. HIV-1 Epidemic Control — Insights from Test-and-Treat Trials. N Engl J Med. 2019 Jul 17;381(3):286–8.

5. Fauci AS, Lane HC. Four decades of HIV/AIDS—much accomplished, much to do. New England Journal of Medicine. 2020;383(1):1–4.

6. United Nations. Addressing inequalities and getting back on track to end AIDS by 2030: Report of the Secretary-General [Internet]. New York: UN; 2020 [cited 2021 May 12]. Report No.: A/75/836. Available from: https://hlm2021aids.unaids.org/sg-report/

7. UNAIDS. Prevailing Against Pandemics [Internet]. Geneva: UNAIDS; 2020 [cited 2021 Jan 4]. Available from: https://aidstargets2025.unaids.org/

8. Global Commission on HIV and the Law. HIV and the Law: Risks, Rights & Health [Internet]. New York: United Nations Development Program; 2012. Available from: https://hivlawcommission.org/

9. UNAIDS. HIV Prevention Among Key Populations [Internet]. Geneva: UNAIDS; 2016 Nov. Available from: http://www.unaids.org/en/resources/presscentre/featurestories/2016/november/20161121_keypops

10. UNAIDS. 90-90-90: An ambitious treatment target to help end the AIDS epidemic [Internet]. 2014 [cited 2020 May 14] p. 40. Available from: https://www.unaids.org/sites/default/files/media_asset/90-90-90_en.pdf

11. UNAIDS. Global AIDS Strategy 2021-2026: End Inequalities, End AIDS [Internet]. 2021 [cited 2021 May 14]. Available from: https://www.unaids.org/en/Global-AIDS-Strategy-2021-2026

12. Kavanagh MM, Graeden E, Pillinger M, Singh R, Eaneff S, Bendaud V, et al. Understanding and comparing HIV-related law and policy environments: cross-national data and accountability for the global AIDS response. BMJ Global Health. 2020;5(9):e003695.

13. Richard A. McKay. Before HIV. In: The Routledge History of Disease [Internet]. Routledge; 2016 [cited 2021 May 15]. Available from: https://www.routledgehandbooks.com/doi/10.4324/9781315543420.ch24

14. Whitehead J. Sir Keir Starmer says he opposes relaxing drugs laws and insists he is “proud” to be patriotic. iNews [Internet]. 2021 Feb 21 [cited 2021 May 13]; Available from: https://inews.co.uk/news/politics/keir-starmer-drug-liberalisation-laws-health-crime-881386

15. Hayes Smith R, Shekarkhar Z. Why is prostitution criminalized? An alternative viewpoint on the construction of sex work. Contemporary Justice Review. 2010;13(1):43–55.

16. Farley M. “Bad for the body, bad for the heart”: Prostitution harms women even if legalized or decriminalized. Violence Against Women. 2004;10(10):1087–125.

17. Makofane K, Beck J, Lubensky M, Ayala G. Homophobic legislation and its impact on human security. African Security Review. 2014;23(2):186–95.

18. Adebisi YA, Alaran AJ, Akinokun RT, Micheal AI, Ilesanmi EB, Lucero-Prisno III DE. Sex workers should not be forgotten in Africa’s COVID-19 response. The American Journal of Tropical Medicine and Hygiene. 2020;103(5):1780–2.

19. Arreola S, Santos G-M, Beck J, Sundararaj M, Wilson PA, Hebert P, et al. Sexual stigma, criminalization, investment, and access to HIV services among men who have sex with men worldwide. AIDS and Behavior. 2015;19(2):227–34.

20. Beyrer C, Baral SD, Collins C, Richardson ET, Sullivan PS, Sanchez J, et al. The global response to HIV in men who have sex with men. The Lancet. 2016;388(10040):198–206.

21. Beyrer C, Crago A-L, Bekker L-G, Butler J, Shannon K, Kerrigan D, et al. An action agenda for HIV and sex workers. The Lancet. 2015;385(9964):287–301.

22. Strathdee SA, Beletsky L, Kerr T. HIV, drugs and the legal environment. International Journal of Drug Policy. 2015;26:S27–32.

23. Trapence G, Collins C, Avrett S, Carr R, Sanchez H, Ayala G, et al. From personal survival to public health: community leadership by men who have sex with men in the response to HIV. The Lancet. 2012;380(9839):400–10.

24. Poteat T, Diouf D, Drame FM, Ndaw M, Traore C, Dhaliwal M, et al. HIV risk among MSM in Senegal: a qualitative rapid assessment of the impact of enforcing laws that criminalize same sex practices. PloS One. 2011;6(12):e28760.

25. Sarang A, Rhodes T, Sheon N. Systemic barriers accessing HIV treatment among people who inject drugs in Russia: a qualitative study. Health Policy and Planning. 2012;czs107.

26. King R, Sebyala Z, Ogwal M, Aluzimbi G, Apondi R, Reynolds S, et al. How men who have sex with men experience HIV health services in Kampala, Uganda. BMJ Global Health. 2020 Apr 1;5(4):e001901.

27. Shannon K, Strathdee SA, Goldenberg SM, Duff P, Mwangi P, Rusakova M, et al. Global epidemiology of HIV among female sex workers: influence of structural determinants. The Lancet. 2015;385(9962):55–71.

28. Reeves A, Steele S, Stuckler D, McKee M, Amato-Gauci A, Semenza JC. National sex work policy and HIV prevalence among sex workers: an ecological regression analysis of 27 European countries. The Lancet HIV. 2017;4(3):e134–40.

29. Lyons C, Diouf D, Rwema JO, Kouanda S, Simplice A, Kouame A, et al. Utilizing individual level data to assess the relationship between prevalent HIV infection and punitive same sex policies and legal barriers across 10 countries in Sub-Saharan Africa. In: Journal of The International AIDS Society. IAS; 2020. p. 102– 102.

30. Lyons CE, Schwartz SR, Murray SM, Shannon K, Diouf D, Mothopeng T, et al. The role of sex work laws and stigmas in increasing HIV risks among sex workers. Nature Communications. 2020;11(1):1–10.

31. Papke LE, Wooldridge JM. Econometric methods for fractional response variables with an application to 401 (k) plan participation rates. Journal of Applied Econometrics. 1996;11(6):619–32.

32. UN Office of the High Commissioner for Human Rights. Principles relating to the Status of National Institutions [Internet]. Geneva: United Nations; 1993 [cited 2021 May 13]. Report No.: UNGA Res 48/134. Available from: https://www.ohchr.org/en/professionalinterest/pages/statusofnationalinstitutions.aspx

33. World Bank. World Development Indicators [Internet]. 2020 [cited 2021 May 14]. Available from: https://databank.worldbank.org/source/world-development-indicators

34. UNAIDS. AIDSinfo [Internet]. Available from: http://aidsinfo.unaids.org/

35. Ross M. Is Democracy Good for the Poor? American Journal of Political Science. 2006;50(4):860–74.

36. Boily M-C, Shannon K. Criminal law, sex work, HIV: need for multi-level research. The Lancet HIV. 2017;4(3):e98–9.

